# Community Perspectives on COVID-19 Vaccine Allocation Ethical Principles in Uganda: A Descriptive Cross-Sectional Study

**DOI:** 10.1101/2025.03.12.25323855

**Authors:** Juliet Kiguli, Stuart Ssebibubbu, Joyce Nabaliisa, Celia Nalwadda, Bob Ramadahan Kirunda, Mooka Kamweri

## Abstract

The COVID-19 pandemic has posed significant challenges globally, necessitating the rapid development and distribution of vaccines. This study aimed to engage community members in Uganda to understand their perceptions of the ethical principles and prioritisation criteria for COVID-19 vaccine allocation. Employing a mixed-method approach of quantitative survey and qualitative key informant interviews, the study yielded significant findings crucial for understanding community perspectives on COVID-19 vaccine allocation in Uganda.

This study investigated Ugandan community perspectives on the ethical principles governing COVID-19 vaccine allocation. While 82% of respondents were familiar with vaccination guidelines, vaccine hesitancy was evident due to mistrust, concerns about information quality, preference for alternative medicine, and doubts regarding vaccine efficacy.

Despite general support for ethical principles, many participants doubted the government’s ability to ensure equitable vaccine distribution. Concerns about fairness and global equity for low-middle-income countries were raised, emphasising the need to address cost, corruption, and political influence.

The study highlights the importance of contextualising and localising ethical principles to enhance their relevance and acceptance within communities. Effective communication, community engagement, and addressing underlying sociocultural and systemic barriers are crucial for successful vaccine deployment. Incorporating community perspectives can foster trust and ownership, ultimately improving vaccine uptake and pandemic response.

## Introduction

The COVID-19 pandemic, a global health crisis of unprecedented scale, has necessitated the rapid development and distribution of vaccines to control the spread of the virus. In December 2019, China reported 44 “Pneumonia of unknown cause” cases to the World Health Organisation(1). Later, in February 2020, this disease that later came to be known as coronavirus disease 2019 (COVID-19) was declared “a Public Health Emergency of International Concern, a call for global solidarity and cooperation in the face of a common threat (2). By July 2020, the impact of the pandemic on Africa was significant, with 369,928 infections and 6974 deaths recorded.

According to the Uganda presidential address in March 2020, around the same time, Uganda had registered 953 cases of infection, 892 recoveries, and no deaths(3). Currently, Uganda has a total of 38,085 cases and a total death of 304(4). As of 31 January, Kenya had 100,773 cases and 1,763 deaths. In response to the COVID-19 pandemic, the Governments of Kenya and Uganda adopted similar public health measures to contain its spread. Some initial measures included refusing to repatriate citizens studying in China, mandatory institutional quarantine, and physical and social distancing(5).

In Uganda, as in many other countries, the ethical principles governing vaccine allocation have become a focal point of discussion. Equity, justice, and community engagement principles are particularly salient in this context, as they directly influence public acceptance and the overall effectiveness of vaccination campaigns. The ethical allocation of vaccines is not merely a logistical challenge but also a moral imperative, especially in resource-limited settings where disparities in healthcare access are pronounced(6),(7).

The concept of beneficence, which emphasises the importance of acting in the best interest of individuals and communities, is central to the ethical discourse surrounding vaccine distribution. Vaccination programs must not only aim to achieve herd immunity but also ensure that vulnerable populations are prioritised(8),(7). In Uganda, where healthcare resources are limited, the challenge lies in balancing the need for rapid vaccine deployment with the ethical obligation to ensure equitable access for all segments of the population, particularly marginalised groups who may face barriers to healthcare(9),(10). The World Health Organization’s (WHO) guidelines on vaccine allocation underscore the need for a values-based framework that prioritises health workers and high-risk populations, reflecting a commitment to equity and justice in public health(6).

Community perspectives are pivotal in shaping the ethical landscape of vaccine allocation. Understanding local communities’ concerns, beliefs, and values regarding vaccination can significantly enhance trust and acceptance of the vaccine. In Uganda, historical factors, including past injustices in healthcare, may contribute to vaccine hesitancy, making it crucial for health authorities to adopt a participatory approach incorporating community voices in decision-making processes.

The WHO Values Sage Framework offers guidance globally on the allocation of COVID-19 vaccines between countries and offers guidance nationally on the prioritisation of groups for vaccination within countries while supply is limited(11). The Framework has been developed to provide a values foundation for SAGE recommendations on priority target groups for specific COVID-19 vaccines at different stages of supply availability. It intends to be a helpful tool for policymakers and expert advisors at the global, regional, and national levels as they make allocation and prioritisation decisions about COVID-19 vaccines.

In addition, the Framework is intended to help all stakeholders, including community and advocacy groups, the general public, health professionals, and other civil society organisations, as they contribute to decisions about how limited supplies of COVID-19 vaccines should be deployed for optimal impact. The Framework addresses only ethical issues regarding allocating and prioritising COVID-19 vaccines.

As such, the Framework articulates the overall goal of COVID-19 vaccine deployment and provides six(11) core principles that should guide the distribution of vaccines.

### Human Well-Being

Protect and promote human well-being, including health, social and economic security, human rights and civil liberties, and child development. The principle further requires that those making vaccine allocation and prioritisation decisions determine what vaccine deployment strategies will best promote and protect all the implicated dimensions of well-being, including strategies for containing transmission, reducing severe disease (including long-term sequelae) and death, or a combination.

### Equal Respect

Recognize and treat all human beings with equal moral status and their interests as deserving of equal moral consideration. The principle that all people are and should be treated as moral equals, entitled to equal respect and consideration of their interests, is enshrined in the Universal Declaration of Human Rights31 and the constitutional documents of many countries. Equal respect is also generally understood to be a foundational principle of ethics and of justice or equity in particular.

### Global Equity

Ensure equity in vaccine access and benefit globally among people in all countries, particularly those in low-and middle-income countries. Countries and territories are primarily responsible for protecting and promoting the well-being and human rights of those living within their borders. It is thus reasonable and appropriate for countries to be concerned with securing sufficient COVID-19 vaccines to meet the needs of their populations.

### National Equity

Ensure equity in vaccine access and benefits within countries for groups experiencing more significant burdens from the COVID-19 pandemic. It is essential to use constrained resources efficiently, especially when the resource is as high-value as vaccines in a devastating pandemic.

### Reciprocity

This plays a crucial role in COVID-19 vaccine allocation. It underscores the importance of acknowledging and rewarding those who bear exceptional risks, often due to their occupations, during a pandemic. This principle should be carefully interpreted to prevent inappropriate claims by individuals or entities with disproportionate power and resources.

### Legitimacy

Make global decisions about vaccine allocation and national decisions about vaccine prioritisation through transparent processes based on shared values, best available scientific evidence, and appropriate representation and input by affected parties. Legitimacy in the context of COVID-19 vaccines and this pandemic refers to the appropriate authority to make recommendations and governing decisions about who gets the vaccine and when. Because different stakeholders, including different countries at the global level and interest groups at the national level, are likely to have different views about vaccine allocation and prioritisation. All concerned must know that the recommendations and decisions emanate from a legitimate body through a legitimate process.

These principles are relevant to ensuring that the allocation process enables equitable access and fair allocation of vaccines, therapeutics, and diagnostics. The main objective of this study was to engage Community members in understanding their perceptions of COVID-19 Vaccine Allocation Ethical Principles in Uganda.

#### Specific objectives

1. To assess community members’ feelings, concerns, attitudes, and agreement with the ethical principles and prioritisation criteria for COVID-19 vaccine distribution.
2. To examine the logic/thinking behind the views of community members towards the ethical principles and prioritisation criteria of COVID-19 vaccination.

## Methods and Materials

We adopted a mixed-method systematic, cross-sectional descriptive study. This included both quantitative and qualitative approaches. This mixed-methods approach is particularly evident in understanding the Community perception of COVID-19 Vaccine Allocation Ethical Principles in Uganda through engaging community members. A closed-ended Likert scale questionnaire was used for the quantitative approach. This was mainly used because of the need to investigate perceptions, motives and feelings. On the other hand, open-ended questions were used with selected study participants, and these questions were embedded and combined with quantitative questions in one data collection instrument. Conversely, qualitative data were collected using key informant interviews (KIIs) and an online survey (telephone interviews). The participants’ recruitment period started from 23^rd^ March 2021 until 23^rd^ May 2021 and verbal informed consent was obtained from all participants following the ethical approval obtained. Data were analysed using thematic content analysis with the help of NVivo 12.0. The Study areas were divided into four cultural regions as follows:

1. Central Uganda Region incorporating the Buganda Central & Buganda South;
2. Eastern Uganda Region incorporating Teso, Busoga, Bukedi, Bugisu, Sebei;
3. Western Uganda incorporating Kigezi, Ankole, Toro and
4. Northern Uganda incorporating Langi, Karamojong, Lugbara and Acholi.

### Study Population and Selection Criteria

The study population was identified through community-based organisations to get a broad representation of the various community members and practical community entry points. These CBOs were cultural, social, health initiatives and economic empowerment CBOs. The recruitment and sampling process was led by the CACVAEP study coordinator, who obtained a list of community-based organisations from the civil society organisation network (CSO) focal person within the region. Later, individuals from the community-based organisations were categorised into four major groups: – women, men, youth and special groups such as people living with Disability, People living with HIV, Traditional healers and village elders. Within each group, study participants were purposively sampled across the various groups to meet the following inclusion criteria:

- Must be the age of 18 years and above;
- Must own a smartphone or laptop
- Must know the national languages
- Give consent to participate in the survey

### Data Analysis

The Statistical Package for Social Sciences (SPSS) was used to analyse the quantitative data. Before analysis, the data was thoroughly reviewed and cleaned to ensure accuracy and consistency. Descriptive statistics summarised the variables, including simple frequencies, proportions, and percentages. To test factors associated with the acceptance of the COVID-19 vaccine, inferential statistics, specifically the chi-square test and bivariate analysis, were employed. A 95% confidence interval and a significance level of p < 0.05 were adopted for statistical significance.

Qualitative data was analysed using NVivo(12). This qualitative analysis was primarily used to complement the quantitative survey and address potential biases that may have arisen from closed-ended or leading questions. Combining quantitative and qualitative data gave a more comprehensive understanding of the factors influencing COVID-19 vaccine acceptance.

### Ethical consideration

The Uganda National Council of Science and Technology’s Ethical Committee approved the study. Before administering the questionnaire, all study participants provided written informed consent. This ensured that participants were fully aware of the study’s purpose, potential risks and benefits, and their right to withdraw at any time.

## Results and Discussion

This was a purposive nonprobability study by design, and 100 respondents referred by local community organisations were interviewed. Analysis of the participants by category shows that most respondents were local community influencers whose voices were respected at the community level but held no particular office. It is important to note that while political and cultural influencers were the least identified, they yield a lot of power of influence on behavioural change and, thus, must not be underestimated. Forty-three per cent (43%) were female, while 57% were male respondents. Figure a above shows participants by category.

**Figure a:**
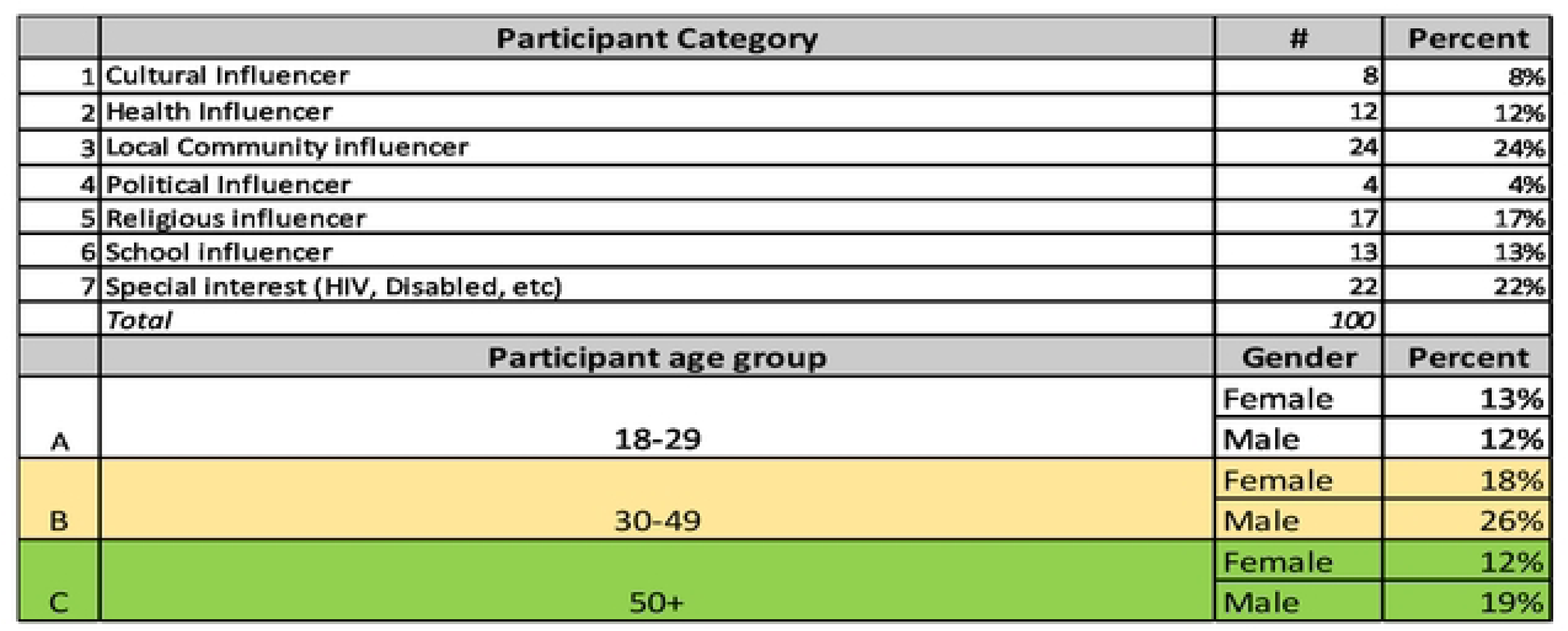
Social Demographics of Respondents

### Section A: Knowledge, awareness and attitude towards COVID-19

This study was based on WHO ethical principles and prioritisation in relation to local community knowledge, beliefs, perceptions, and feelings. To this extent, we sought to determine respondents’ knowledge of WHO and National/MOH COVID-19 guidelines. Results showed that 82% were aware of either WHO or National/MOH guidelines on COVID19 vaccination. However, 15% were unaware of either WHO or MOH guidelines on vaccination, pointing to a communication gap. It should be noted that 36% and 46% were aware of WHO and National guidelines, respectively, suggesting that a uniform global and local/National communication plan for COVID-19 and most likely other pandemics is vital. Otherwise, if either WHO or MOH were not, 36% or 46% would most likely be unaware of COVID-19 vaccination guidelines. Table 1 shows the knowledge and awareness by source.

**Table 1.** Awareness of WHO and Uganda MOH COVID19 vaccination guidelines.

| Awareness of WHO and National/MOH COVID guidelines | Percent |
| --- | --- |
| Know both WHO and Uganda/MOH guidelines | 36% |
| Know Uganda/MOH guidelines only | 46% |
| Know WHO guidelines only | 3% |
| Don't know either WHO or Uganda MOH guidelines | 15% |

Several studies indicate that vaccination is vital for protection and can potentially reduce the transmission of infectious diseases. Thus, this study inquired into community levels of agreement that the COVID-19 vaccine can curb the spread of the pandemic in Uganda.

Findings revealed that only 35% of respondents agreed/strongly agreed that the COVID-19 vaccine can curb the spread of the pandemic. Sixty-five per cent (65%) were either neutral/somewhat agree or disagree/strongly disagree. Specifically, 23% disagreed/strongly disagreed, while the most significant proportion (42%) were neutral/somewhat agreed, with strong doubts about the effectiveness of vaccination in curbing the spread of COVID-19.

Thematic analysis of reasons behind those who disagreed revealed that a) Mistrust, b) Information quality, c) alternative medicine, d) Vaccine effectiveness and e) mixed government signals are vital in influencing people’s attitudes and behaviour/actions towards the COVID-19 vaccine. Therefore, any response to positively influencing communities to accept and trust that the vaccine will curb the spread of the pandemic must address the fears and concerns around the five themes identified above. The same themes emerged from data analysis from respondents who were neutral/somewhat agreed (sitting on the fence). Figure b below shows the respondents’ level of agreement and reasoning.

**Figure b:**
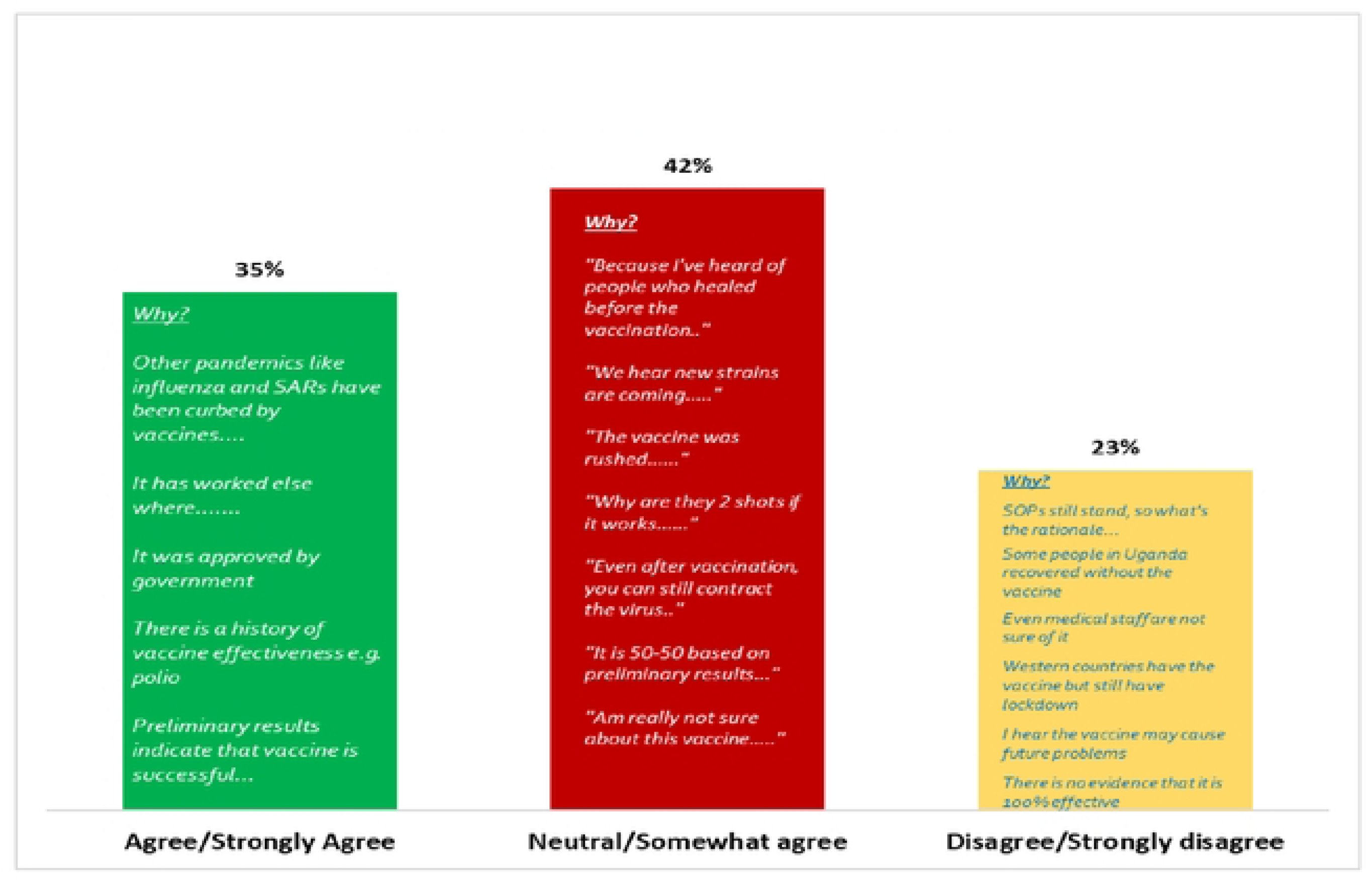
Level of agreement that vaccination can curb the spread of COVID-19 in Uganda

### Section B: Community feelings, concerns, attitudes and agreement with/around the ethical principles and prioritisation criteria for COVID-19 vaccine distribution

Commitment to equity across all countries and for all populations in need remains the foundation for a global fair allocation framework(13). Fairly distributing a COVID-19 vaccine among countries is a problem of distributive justice. Although governments will be the initial recipients of vaccines, fair distribution across countries must reflect a moral concern for the ultimate recipients, who are individuals. Three values are particularly relevant: benefiting people and limiting harm, prioritising the disadvantaged and equal moral concern(13). The ethical principles that/around which the study inquired and the findings are elaborated below.

#### Human Well-Being

This principle emphasises protecting and promoting human well-being, including health, social and economic security, human rights and civil liberties, and child development. Findings showed that 94% agreed with this principle, with 81% saying that the government should get the vaccine for everyone. We also asked about their likelihood of acquiring the COVID-19 vaccine even after getting the vaccine, and results showed that 58% of the community noted that they are likely to acquire the virus even with the vaccine. This means that the vaccine will, in fact, not guarantee human protection and well-being as envisaged in principle, thus raising the question: How can the principle of Human well-being be achieved in the expanding COVID-19 vaccination? Is there an alternative way of attaining this principle?

Findings show that 15% believe it is not likely for them to contract COVID-19 once they get the vaccine, which is contrary to what the scientific evidence says. Another 27% do not know if they are likely to contract the virus again after being vaccinated. These suggest a gap in the communication strategy, approach and information quality about how the COVID-19 vaccine works—especially on its level of effectiveness. Figure c below shows the level of likelihood of acquiring COVID-19 after vaccination.

**Figure c:**
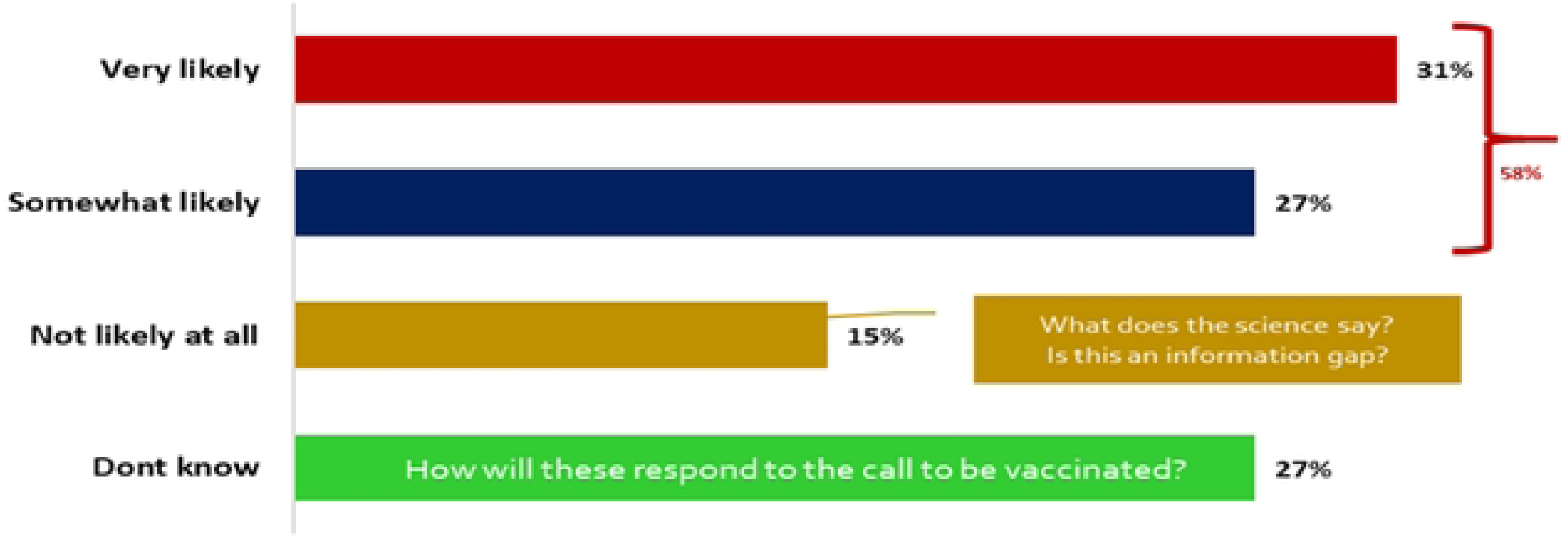
Likelihood of contracting COVID-19 after vaccination

#### Equal Respect

This principle recognises and treats all human beings as having equal moral status and interests, as 81% agreed/strongly agreed that vaccination should treat all people equally, but only 60% trust that the government /MOH will ensure that all people are treated equally. Only 8% feel that vaccination will not discriminate against social class. Thus, while the community agrees with the principle of equality, they don’t think or trust that this principle is achievable in Uganda. The most frequently mentioned factors likely to affect equality in vaccine distribution are limited accessibility, age discrimination, religious biases, social class/status, corruption, tribalism and politicisation, as listed in Table 2.

**Table 2:**
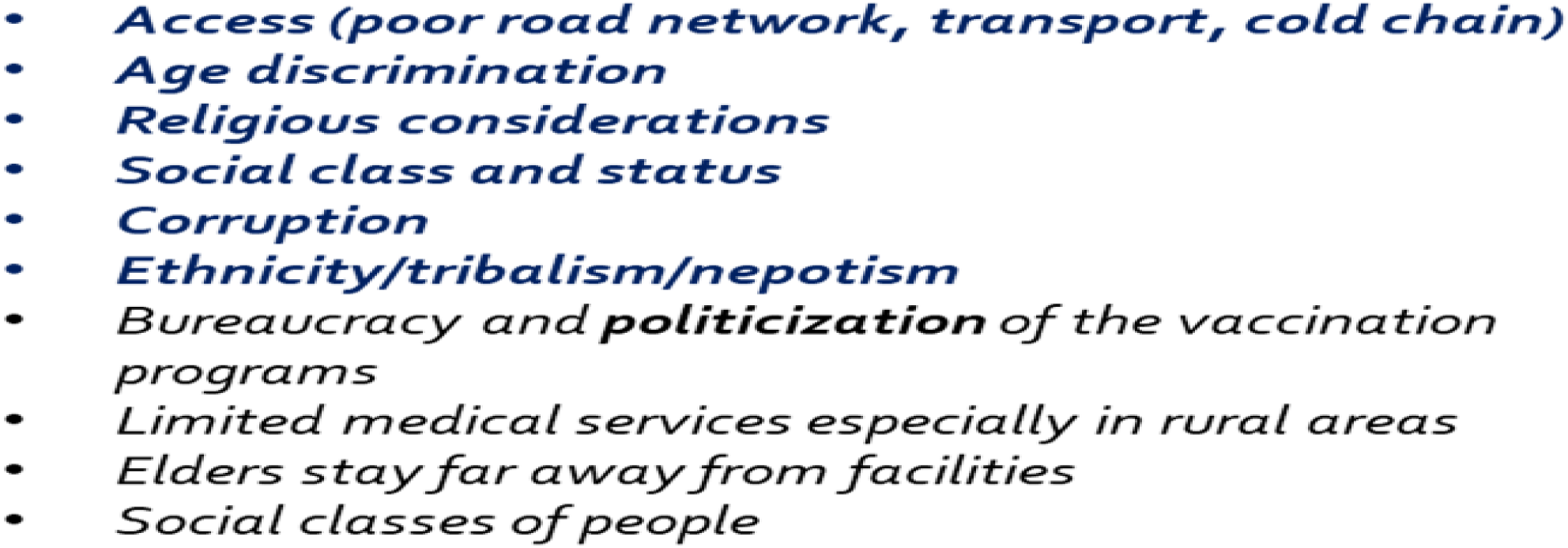
Factors that may affect equality in vaccine distribution.

Figure d below illustrates their feelings, trust, and perceptions. Therefore, the government needs to ensure that the vaccination program addresses fears if the community is to trust vaccine distribution for better vaccine uptake.

**Figure d:**
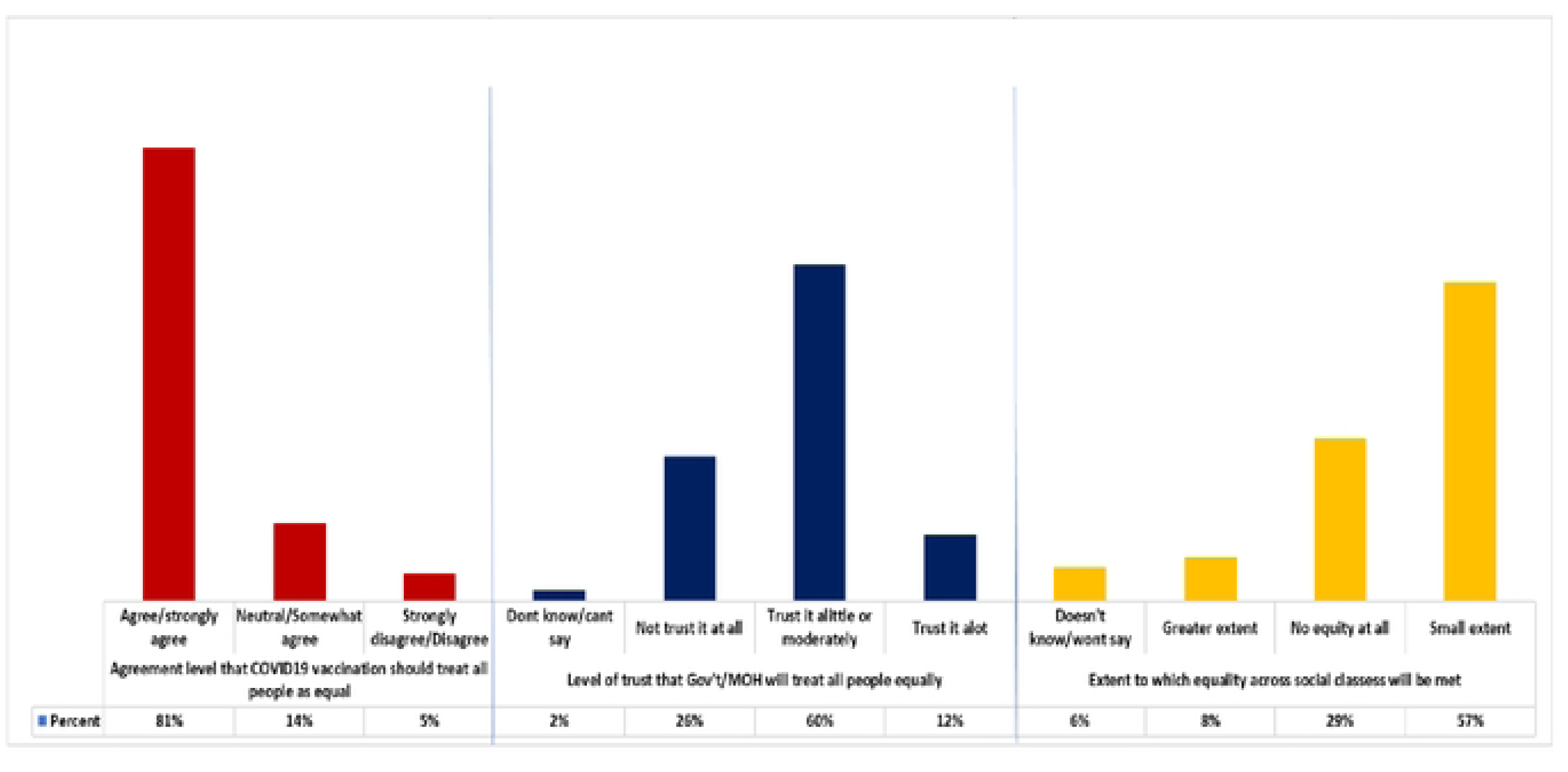
Community feelings, level of trust and perceptions around the equality principle

The study sought to understand the influence of social-cultural constructs, beliefs and equality in vaccine distribution and access. Findings from the content analysis show that factors were around the following themes:

- **Gender and power** (superiority of men versus women): The study found that women and children may access vaccines later after men get their vaccines. Where men are against the vaccine, women may be prohibited, especially if their husbands are against it. Respondents reported that men have the upper hand over women. Therefore, it is essential for the vaccination distribution program to consider gender dynamics and power if the equality principle is to be achieved as envisaged.
- **Economic status**: Society is constructed along the lines of poor—rich. Thus, the poor think they can only access the vaccine after the rich receive their shots. This attitude may affect the pace of distribution, with the thinking that the poor majority come last.
- **Social norms influence**: Negative social norms are greatly responsible for hesitancy about immunisation programs, and this is no different for the COVID-19 vaccine. Results indicate that social norms around vaccination still prevail. However, to ensure successful vaccine distribution in Uganda, it is necessary to explore particular norms and their drivers.
- **Herbal alternatives: The community** believes that Uganda has not had massive deaths because Ugandans have been using herbal mixtures, and they work. Thus, we will likely see low uptake in rural communities where herbal medicine is more frequently used due to other factors, including poor access to health services.
- **Comparative supremacy in tribal identity:** The results suggest that Ugandan society is rapidly dividing along tribal lines. A significant portion of community members now believes that they are either superior or inferior to other tribes and thus are entitled or less/not entitled to services—in this case, the COVID-19 vaccine. Urgent and deliberate efforts must be undertaken to deconstruct these beliefs.

#### Global Equity

This principle ensures equity in vaccine access and benefits globally among people in all countries, particularly those in low-and middle-income countries. Findings around this principle show that 76% of the community respondents agree that vaccination distribution should ensure global equity. Eighty-two per cent (82%) of the community believe that it’s not likely (41%), somewhat likely (41%), or they don’t know (9%) if the vaccine distribution will ensure equity for low and middle-income countries as illustrated in figure e.

**Figure e:**
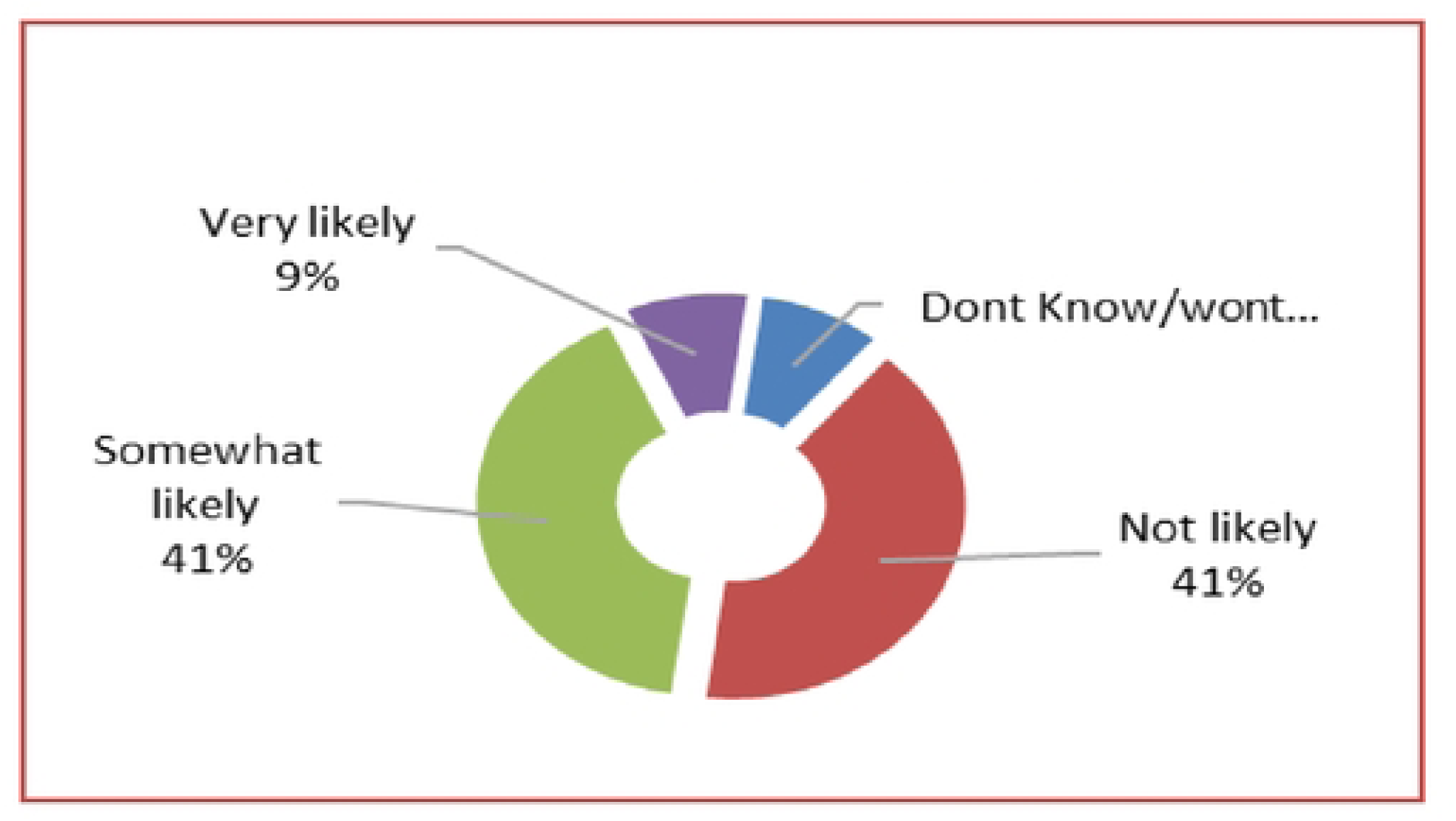
Likelihood that vaccine distribution will ensure equity for LMICs

The most highlighted reasons include:

- The cost of the vaccine is too high
- Poor systems for vaccine distribution
- Since LMICs are poor, they have no choice
- Globally, we have received different vaccines already, so there can’t be equity
- Corruption in African countries may affect WHO’s effort
- Only a few countries have been able to receive the vaccines so far
- Richer countries have the vaccine more readily accessible to them than LMICs
- COVID-19 is more of a political risk than a health risk….
- Already, few doses were brought for the prioritised groups and not everyone

In terms of community perception and feelings around vaccine quality, the findings, as illustrated in Figure f, revealed that 87% of respondents said they either don’t trust at all (57%) or trust a little/moderately (30%) that LMICs and HICs will get the same quality of vaccine. They noted that already, there are types of vaccines that are not accessible to African countries, the excellent quality vaccines are expensive, LMICs rely on donations and have no choice, and the vaccines are expensive. These resonate with the reasons put forward by communities as being in the way of achieving global equity. Results also show that the Ugandan community doesn’t fully trust that global agencies will meet Uganda’s vaccine needs.

**Figure f:**
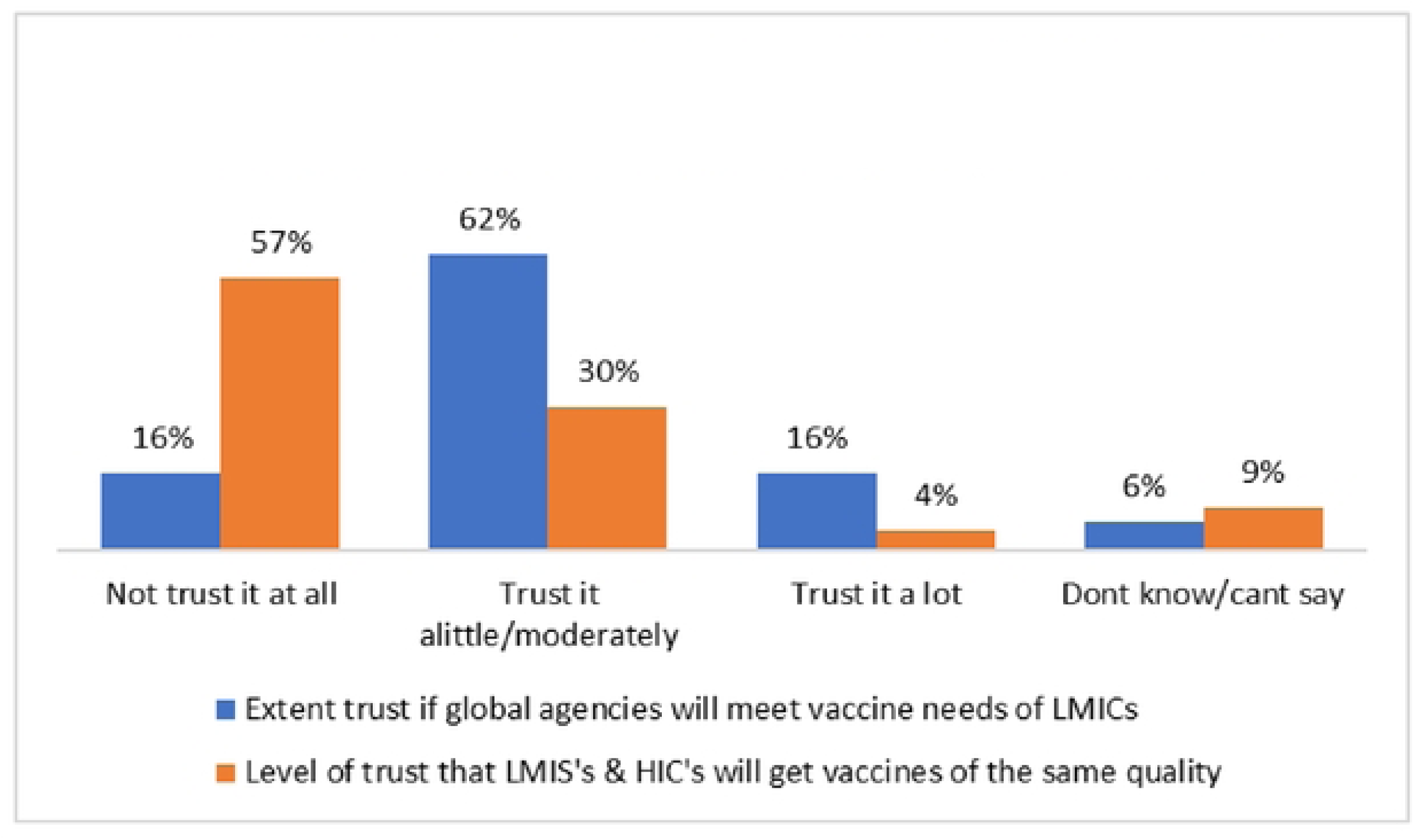
Trust that global agencies including WHO will meet vaccine needs and quality for LMICs

In the spirit of “potential co-production,” the study sought to hear from the community strategies for ensuring that Uganda gets vaccines of the same quality. Table 3 below shows the community suggestions that critical stakeholders could adapt for more effective and community-centred vaccine distribution.

**Table 3:** Strategies to ensure that we get vaccines of the same quality.

|  |
| --- |
| **Table 3: Strategies to ensure that we get vaccines of the same quality** |
| *1 - Get Vaccines from trusted countries* |
| *2 - Ugandan Researchers test vaccines* |
| *3 - WHO should ensure the vaccine is from one supplier* |
| *4 - WHO should guide on the vaccines given out* |
| *5 - Quality control by WHO before allowing vaccines on the market* |

Once the community is not sure about the quality of the vaccine, sees that mortality is not as bad as elsewhere, and is presented with alternatives, including herbal mixtures, vaccine hesitancy becomes almost inevitable. Global quality should thus not be questioned if the COVID-19 vaccine distribution is to be successful.

#### National Equity

This principle aims at ensuring equity in vaccine access and benefits within countries for groups experiencing more significant burdens from the COVID-19 pandemic. Results show that 86% of the community agrees/strongly agree that vaccine distribution should strongly consider National Equity. However, 85% either don’t trust at all (22%) or trust a little/moderately (63%) that we shall have equity in access to the COVID-19 vaccine in Uganda. The top-ranked reasons include corruption, nepotism/tribalism, weak systems, discrimination (class and geography) and lack of total government commitment, among others.

#### Reciprocity

This is about honouring obligations of reciprocity to those individuals and groups within countries who bear significant additional risks and burdens of COVID-19 response for the benefit of society. Results show that 62% of the respondents agree that vaccination should prioritise those with additional risk. Still, 74% do not think that it is likely that the vaccine distribution will reach those with additional risk faster. This speaks to the nature of Uganda’s immunisation capacity, corruption, mistrust, and discrimination, which were reported earlier.

#### Legitimacy

This principle aims to make global decisions about vaccine allocation and national decisions about vaccine prioritisation through transparent processes based on shared values, best available scientific evidence, appropriate representation, and input by affected parties. Study findings (figure g) showed that 77% of the community members don’t trust at all (24) or trust a little (53%) that Uganda was engaged in decision-making. This lack of trust and perception could explain why most community members don’t trust the quality of vaccines that Uganda receives and distributes. Broadly, it could also speak to “Public-Health Power” relations between the LMICs and HICs where vaccines are manufactured.

**Figure g:**
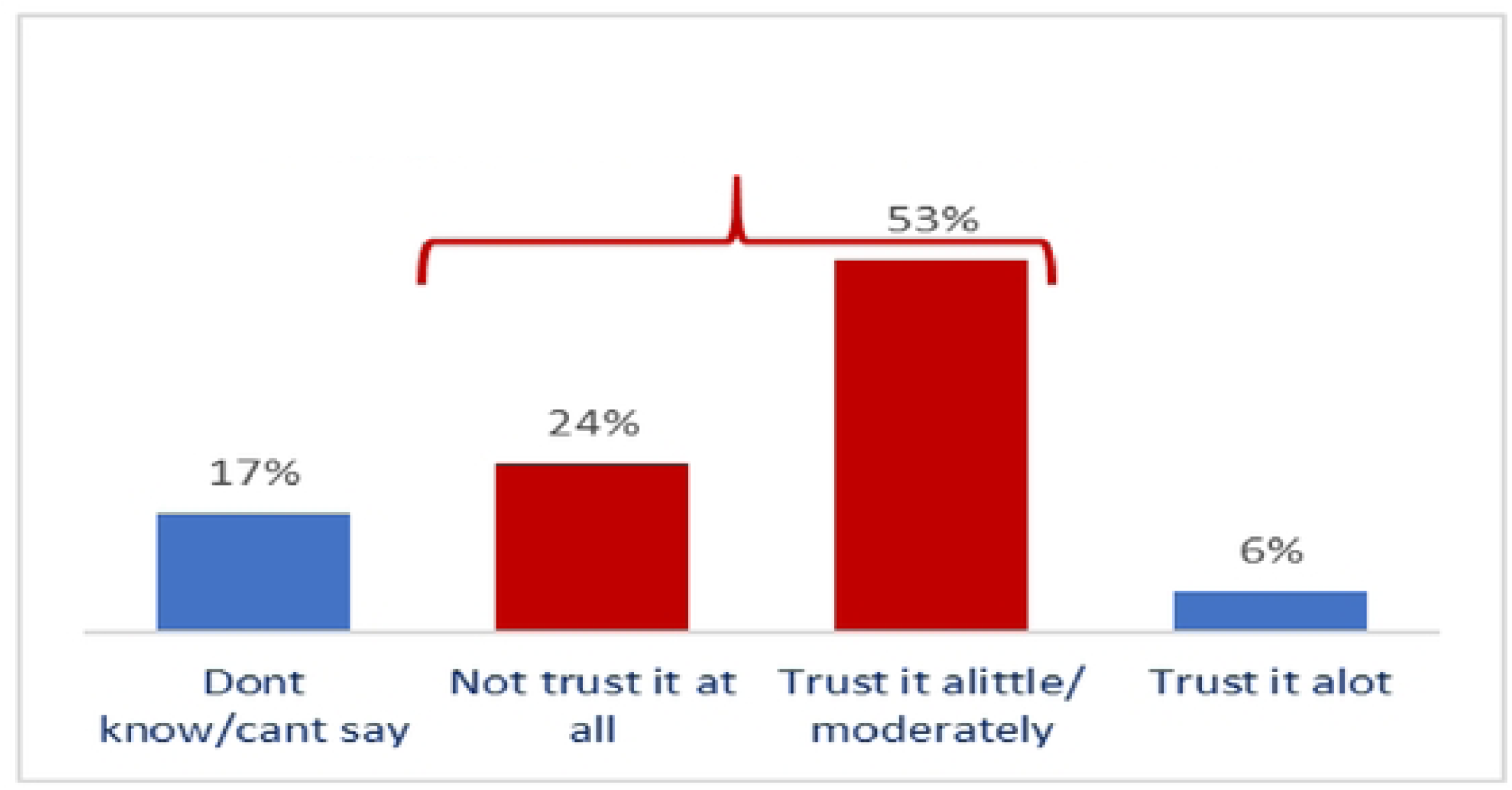
Level of trust that Uganda was engaged in COVID 19 vaccine decisions

**Figure h:** Extent of agreement that vaccines will be fairly distributed to individual/regions

In terms of distributing vaccines fairly to individuals and regions, 77% of the community either disagrees/strongly disagrees (31%) or is neutral/somewhat agrees (46%) that vaccine distribution will be fair. If not addressed, these feelings and perceptions will derail Uganda’s COVID-19 vaccination campaign.

Therefore, if this principle is to make practical local sense to the target user of the service/vaccine, it is better to design strategies that move 77% of the community from the undesired red zone into the green zone. Strategies can be at the Global and National levels but with community reach.

The study also sought out who, in the community’s opinion, was left out of the COVID-19 vaccination distribution campaign and process. Results in the word map below indicate that religious leaders, cultural leaders, traditional herbalists/healers, youths, celebrities and local councils were left out. It should be noted that programs that face perception, norm and belief-based barriers, including vaccination programs, are more likely to succeed when influencers with non-enforceable but more powerful power than the power of enforcers are brought on board.

Regarding demand and supply of COVID-19 vaccine, 57% think that demand will outstrip the supply of commodities. These are very concerned about this possibility, noting that more people will die, the spread will increase, the health ministry will be overwhelmed, and the lockdown will be re-instated. Furthermore, 96% believe that accountability for COVID-19 vaccine distribution is essential, but 69% do not trust that Uganda will be accountable. The top tanked reasons they gave were: a) Corruption in government, b) Political influence of the vaccination, c) Previous precedent of lack of vaccine accountability, and d) Poor documentation and monitoring systems. This finding points to a need to be intentional in accountability from a systems point of view.

#### Theoretical Interpretation

Drawing from the lessons learned from other allocation frameworks, the study concludes with an in-depth description and community understanding of the ethical principles of COVID-19 vaccine allocation. The study presents community perceptions and decisions about COVID-19 with uncertainty about the selection criteria. These unknowns include the safety and efficacy of the vaccines in specific populations (such as children, pregnant women, older adults, and individuals previously infected with COVID-19); the effectiveness of vaccines in tandem with existing preventive measures; public confidence in the vaccine; the possibility of ultra-cold storage requirements for the vaccine; the pharmacovigilance evidence; and many other unknowns.

## Conclusion

Identifying the most vulnerable groups in the pandemic so that they may be prioritised in vaccine distribution can help shape our response within one country and globally. Vaccine nationalism poses a significant threat to eradicating COVID-19 by undermining the recognition of all people’s dignity and right to health beyond national borders. In Uganda, despite the good intentions, depth and breadth covered by the ethical principles and prioritisation criteria for COVID-19 vaccines, there was no intentional effort to localise them to ease adaptation. Uganda’s MOH adopted ethical guidelines that were not contextualised and tested before roll-out.

All WHO principles are relevant for ensuring that the allocation process enables equitable access and fair allocation of COVID-19 vaccines. However, for these principles to be practical, they should be localised and contextualised. If communicated well and accurately, they potentially reduce or avert vaccine hesitancy. The study identified several contradictions between what the principles are designed to achieve and facts on the ground/community level. These principles must be further harmonised; otherwise, COVID-19 vaccine hesitancy could be inevitable.

## Data Availability

All materials used for preparation of this manuscript are available upon request from the corresponding author

## Acknowledgements

Not applicable.

## Author Contributions

SS; JN; Conceptualization, design, manuscript writing, and editing. JK; was the PI and led the entire study. All authors contributed to manuscript writing and proofreading and approved the final manuscript.

## Ethics approval and consent to participate

Ethical Approval was granted by the Uganda National Council of Science and Technology (UNCST) through the Makerere University School of Health Sciences ethical committee, Approval No. MAKSHSREC −2021-92. Verbal consent was obtained from all participants in the KIIs, and all participants who filled the questionnaire gave their consent.

## Consent for publication

Not applicable.

